# Oxytocin reshapes fear control: intensity-dependent prefrontal dominance and whole-brain integration during naturalistic viewing

**DOI:** 10.64898/2026.08.11.26360155

**Authors:** Kun Fu, Shuyue Xu, Dan Liu, Zheng Zhang, Qi Liu, Jingxian He, Ting Xu, Can Liu, Junjie Wang, Yuan Zhang, Feng Zhou, Xiaodong Zhang, Chunmei Lan, Mengfan Han, Menghan Li, Zhen Liang, Bharat Biswal, Keith M Kendrick, Weihua Zhao, Dezhong Yao, Benjamin Becker

## Abstract

Anxiety and maladaptive fear remain difficult to treat, and despite initial promising findings, evidence regarding the anxiolytic and translational potential of oxytocin (OT) remains inconsistent. In a preregistered, randomized, double-blind, placebo-controlled, parallel-group pharmaco-fMRI study in 67 healthy men, we tested whether intranasal OT reduces post-exposure subjective fear during prolonged naturalistic viewing a horror movie comprising independently defined low-, medium-, and high-fear segments. OT reduced subjective fear following naturalistic threat exposure after accounting for pre-exposure baseline ratings. Neuroimaging analyses revealed that OT attenuated recruitment of the dorsolateral prefrontal cortex particularly during higher fear, and enhanced coupling of this region with the bilateral amygdala. At the large-scale network level, OT increased communication between frontoparietal/default-mode control networks and subcortical/limbic networks during high fear indicating more integrative fear regulation. A whole-brain fear neuromarker (CAFE) further confirmed intensity-dependent OT effects. Together, these findings indicate that OT modulates post-exposure fear experience and fear-related neural dynamics in ecologically valid contexts.

**Highlights:**

1. Intranasal OT reduced post-exposure subjective fear after controlling for baseline ratings.
2. OT modulated dlPFC activity and dlPFC-amygdala coupling under higher fear contexts.
3. OT increased control-to-limbic/subcortical network communication selectively when fear is elvated.
4. OT altered whole-brain fear-related neural representations in an intensity-dependent manner.

## Introduction

Fear represents an evolutionarily conserved defensive response. Moderate fear and accompanying physiological arousal are crucial for human survival.^1,2^ Fear and anxiety are closely related but distinct affective constructs. Fear is typically elicited by an identifiable and imminent threat, whereas anxiety is more often characterized by sustained anticipation of uncertain or potential future threat.^3,4^ However, excessive and prolonged fear represent a hallmark of mental disorders such as anxiety disorders and post-traumatic stress disorder (PTSD)^5–8^ and are associated with strong functional impairments. As such, these disorders have become a leading cause of disability associated with both personal suffering and socio-economic burden.^9,10^ Accordingly, there is a pressing need for effective strategies to mitigate maladaptive threat-related fear responses, thereby reducing disability and improving clinical outcomes in anxiety disorders and PTSD.

Pathological fear and anxiety-related symptoms are is currently treated by pharmacotherapy, non-invasive brain stimulation (NIBS) and psychotherapy. Pharmacological interventions that target the classical neurotransmitter systems, such as the GABAergic and serotonergic systems, are characterised by moderate response rates and the potential for adverse side effects.^10–12^ Although NIBS offers a promising circuit-level intervention approach, its mechanistic interpretation can be challenging because stimulation effects depend on target site, protocol, baseline brain state, and downstream network engagement,^13–16^ moreover, standard non-invasive stimulation cannot directly target key deep fear-related structures, such as the amygdala, and effects on these regions are typically inferred through connected cortical–subcortical networks.^17,18^ Psychotherapy often requires a high time-investment from patients and application is limited by the lack of qualified therapists.^19,20^ Finding novel, safe and rapid acting approaches to modulate maladaptive fear responses is crucial. Although several potentially efficacious candidate compounds have demonstrated effects in animal models, it is notable that no new pharmacological treatments for fear- and anxiety-related conditions have been introduced into clinical practice over the past several decades.^21–23^

Promising results have been observed in both animal models and preclinical human studies that suggest that neuropeptides, including oxytocin (OT), vasopressin and angiotensin II, may represent potentially effective targets to regulate fear and anxiety.^24–32^ A substantial proportion of literature in this domain has focused on the hypothalamic neuropeptide OT. Animal models^27,28^ and OT administartion studies in humans^26,29–32^ have indicated that OT has the capacity to regulate negative emotional processing in the domains of fear and anxiety.

The potential to regulate fear and anxiety is further underscored by the archieture of the OT system, such that the OT neurons originating from the hypothalamus extend their projections to a multitude of of brain systems that contribute to these emotional states, including the amygdala, hippocampus, midbrain and frontal regions.^1,2,33^ According to the findings of earlier research, intranasal OT has the capacity to modulate these processes by exerting an influence on activity in the amygdala, dorsal anterior cingulate gyrus, and medial prefrontal lobe.^11,29–32,34–37^ It is important to note that, despite the initial promise demonstrated by these studies, the effects of OT on fear and anxiety reduction have been primarily validated in static laboratory paradigms. However, its efficacy under dynamic, naturalistic conditions-which more closely approximate real-life social contexts-remains largely unknown, particularly in light of research indicating that emotional processing differs significantly between these settings.^37–39^ Experiencing fear in real life is contingent upon dynamic interactions with a changing environment and the intensity of the fear experiences fluctuates rapidly in the context of dynamic, real-life situations. In our previous study, we demonstrated that a single intranasal OT dose was sufficient to reduce subjective fear in short naturalistic social contexts.^40^ However, the question of whether these effects will extend to changing fear intensity in longer, dynamic naturalistic contexts remains unknown.

This dynamic perspective is consistent with contemporary models proposing that fear arises from ongoing interactions between subjective appraisal and changes in the external environment, rather than from isolated static stimulus features alone. Therefore, dynamic naturalistic paradigms may be particularly useful for examining how OT modulates fear as it fluctuates over time.^37,38^ Furthermore, these mental processes are underpinned by a sophisticated network of interconnected yet distinct brain systems.^41–43^ To address this complexity and capture the underlying network-level communication during subjective affective experiences, recent conceptual frameworks have advocated for the integration of highly immersive, dynamic movie stimuli with network-level analyses.^44,45^ Building upon this approach, we employed a dynamic rating paradigm to decode movie clips of varying fear intensities, aiming to more accurately investigate the anxiolytic effects of oxytocin in naturalistic settings. This method enables the continuous recording of subjective emotional states (e.g., fear, anxiety, and arousal), thereby establishing a foundation for exploring the mechanisms of emotion processing in complex, dynamic situations.^37,39,46^

It has been demonstrated that fear-related disorders, such as anxiety and PTSD, are associated with not only the hyper- and hypo-activation of specific brain regions, but also with a disruption in the functional connectivity of the wider neural network.^42,47,48^ There is an emerging corpus of research that has begun to elucidate the potential of OT to regulate neurofunctional network-level interactions within fear-related networks in various species.^49–52^ In consideration of the aforementioned findings, it can be hypothesised that these results are, at least in part, attributable to the physiological characteristics inherent to the oxytocin signalling system. These characteristics include axonal release, extensive receptor distribution, and a substantially longer half-life when compared to conventional neurotransmitters.^53^ Consequently, network-level approaches are imperative for comprehensively ascertaining the regulatory influence of OT on the fear experience and its therapeutic potential.

Againt this background the present study employed a pre-registered, randomised, double-blind, placebo-controlled, parallel-group pharmaco-functional MRI (pharmaco-fMRI) design, with n=67 male participants, to capitalise on recent advances in naturalistic fMRI and network-level analyses.

Treatment group (OT vs. PLC) was manipulated as a between-subjects factor, whereas timepoint and fear intensity were modeled as within-subject factors. The participants were asked to attentionally watch a 10-minute horror movie with sound and rate their subjective state before the movie and subjective fear immediatedly after the movie. Based on mean continuous ratings from independent samples (n = 20, 10 males, **Fig 1C**), the movie was divided into low-, medium-, and high-fear segments. Accordingly, we hypothesized that OT would reduce post-exposure subjective fear after accounting for baseline ratings. At the neural level, we tested whether OT modulated fear-intensity-related responses within prefrontal-amygdala circuitry and large-scale networks. Specifically, we expected OT to attenuate fear-related prefrontal recruitment (particularly in the dorsolateral prefrontal cortex, dlPFC, given its established role in initiating top-down regulatory control over amygdala responses during explicit threat processing),^54,55^ alter dlPFC-amygdala functional coupling, and increase integration between higher-order regulatory networks and limbic/subcortical systems during elevated fear. These hypotheses were tested with predefined fear-intensity contrasts and network-level analyses while distinguishing primary inferential analyses from exploratory and descriptive follow-up analyses.

**Fig. 1.**
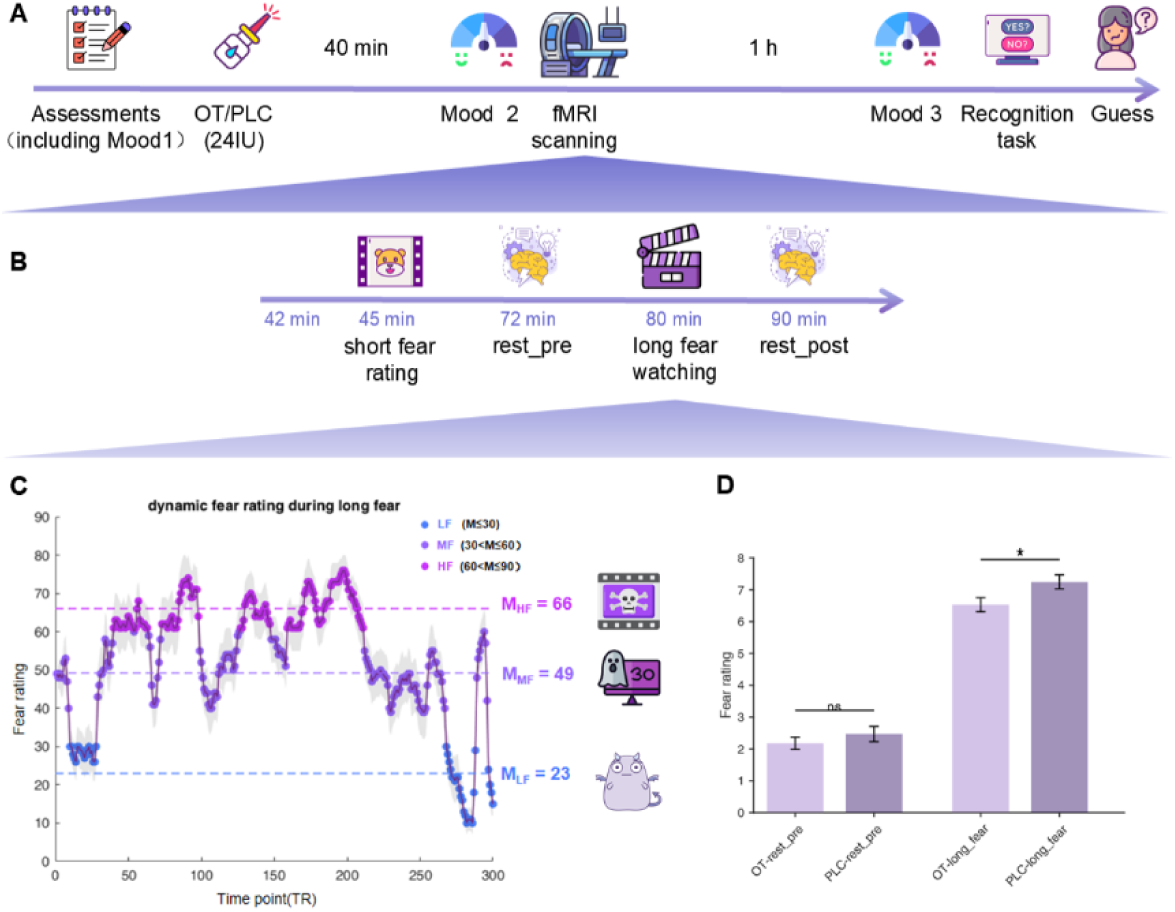
Experimental timeline, paradigm and behavioral fear rating change within and across fMRI sessions. **A** The timeline of the whole experiment. **B** The timeline of sessions during fMRI scanning (The Long Fear task started at 80 min after treatment administration). **C** Dynamic fear rating during Naturalistic fear movie watching paradigm rated by independent sample. **D** The effects of Oxytocin on subjective fear change across fMRI sessions. Abbreviations: OT oxytocin, PLC placebo, fMRI functional magnetic resonance imaging, LF low fear, MF medium fear, HF high fear. License: Images in 1A, 1B and 1C were obtained from https://Flaticon.com under the free license with attribution. * *p*<0.05

## Results

### Demographics and potential confounders

There was no significant difference between the OT (n=33) and PLC (n=34) groups in terms of socio-demographics, mental health indices, or pre-treatment and post-treatment mood assessments arguing against pre-treatment group differences and non-specific treatment effects (**Table 1**, all ps>0.07).

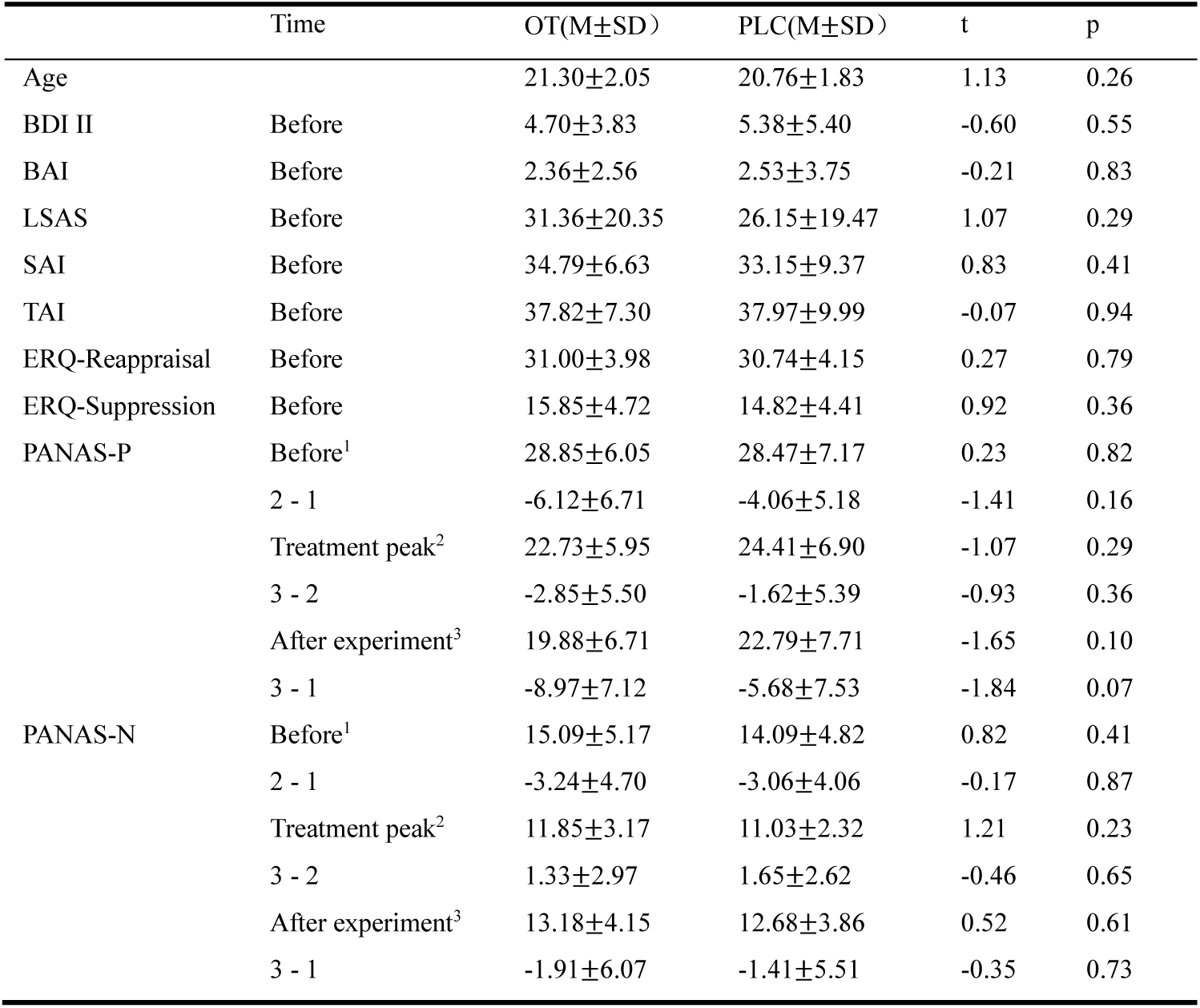
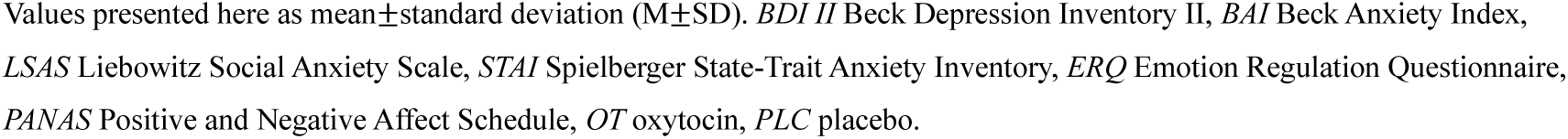

| | Time | OT(M±SD) | PLC(M±SD) | t | p |
| --- | --- | --- | --- | --- | --- |
| Age | | 21.30±2.05 | 20.76±1.83 | 1.13 | 0.26 |
| BDI II | Before | 4.70±3.83 | 5.38±5.40 | -0.60 | 0.55 |
| BAI | Before | 2.36±2.56 | 2.53±3.75 | -0.21 | 0.83 |
| LSAS | Before | 31.36±20.35 | 26.15±19.47 | 1.07 | 0.29 |
| SAI | Before | 34.79±6.63 | 33.15±9.37 | 0.83 | 0.41 |
| TAI | Before | 37.82±7.30 | 37.97±9.99 | -0.07 | 0.94 |
| ERQ-Reappraisal | Before | 31.00±3.98 | 30.74±4.15 | 0.27 | 0.79 |
| ERQ-Suppression | Before | 15.85±4.72 | 14.82±4.41 | 0.92 | 0.36 |
| PANAS-P | Before<sup>1</sup> | 28.85±6.05 | 28.47±7.17 | 0.23 | 0.82 |
| | 2 - 1 | -6.12±6.71 | -4.06±5.18 | -1.41 | 0.16 |
| | Treatment peak<sup>2</sup> | 22.73±5.95 | 24.41±6.90 | -1.07 | 0.29 |
| | 3 - 2 | -2.85±5.50 | -1.62±5.39 | -0.93 | 0.36 |
| | After experiment<sup>3</sup> | 19.88±6.71 | 22.79±7.71 | -1.65 | 0.10 |
| | 3 - 1 | -8.97±7.12 | -5.68±7.53 | -1.84 | 0.07 |
| PANAS-N | Before<sup>1</sup> | 15.09±5.17 | 14.09±4.82 | 0.82 | 0.41 |
| | 2 - 1 | -3.24±4.70 | -3.06±4.06 | -0.17 | 0.87 |
| | Treatment peak<sup>2</sup> | 11.85±3.17 | 11.03±2.32 | 1.21 | 0.23 |
| | 3 - 2 | 1.33±2.97 | 1.65±2.62 | -0.46 | 0.65 |
| | After experiment<sup>3</sup> | 13.18±4.15 | 12.68±3.86 | 0.52 | 0.61 |
| | 3 - 1 | -1.91±6.07 | -1.41±5.51 | -0.35 | 0.73 |

Values presented here as mean±standard deviation (M±SD). *BDI II* Beck Depression Inventory II, *BAI* Beck Anxiety Index, *LSAS* Liebowitz Social Anxiety Scale, *STAI* Spielberger State-Trait Anxiety Inventory, *ERQ* Emotion Regulation Questionnaire, *PANAS* Positive and Negative Affect Schedule, *OT* oxytocin, *PLC* placebo.

### OT reduced subjective fear experience in dynamic changing naturalistic contexts

Effects of OT on subjective fear changes were examined using a mixed repeated-measures ANOVA with timepoint (pre-exposure baseline [Rest_pre] vs. post-exposure [Long Fear]) and treatment (OT vs. PLC) as factors. The analysis revealed a significant main effect of timepoint (*F*_(1,65)_=612.98, *p*<0.0001,*η*_p_^2^=0.904), confirming that fear ratings significantly increased from baseline to the Long Fear phase in both groups. A significant main effect of treatment was also observed, (*F*_(1,65)_=4.66, *p*=0.035,*η*_p_^2^=0.067) indicating overall lower fear ratings in the OT group compared with the PLC group. The interaction effect was not significant (*F*_(1,65)_=1.99, *p* =0.163, *η*_p_²=0.030). Exploratory follow-up comparisons showed no group difference at pre-exposure baseline (*t*_(1,65)_=-0.93, *p*=0.357, Cohen’s *d*=-0.23, **Fig. 1D**), whereas the OT group reported lower post-exposure Long Fear ratings than the PLC group (*t*_(1,65)_=-2.55, *p*=0.013, Cohen’s *d*=-0.62, **Fig. 1D**).

To further control for potential baseline-related variance, an ANCOVA was conducted with long fear rating as the dependent variable, treatment as the between-subject factor, and baseline rating as the covariate. The homogeneity of regression slopes assumption was satisfied, *p* = 0.210. Importantly, after controlling for baseline fear ratings, the OT group showed significantly lower fear ratings than the PLC group following long fear, *β* = −0.718, SE = 0.306, *t*_(1,64)_ = −2.35, *p* = 0.022, *η*_p_² = 0.079. The adjusted Long Fear mean was 7.25 for the PLC group and 6.53 for the OT group. Together, these results indicate successful fear induction by the long horror movie and suggest that OT reduced subjective fear during the Long Fear phase in a highly dynamic naturalistic environment, particularly after accounting for baseline fear ratings.

### OT specifically decreased dorsolateral prefrontal activation in high fear contexts

To explore the brain regions coding levels of subjective fear in naturalistic contexts we first computed the whole-brain activation maps for the fear levels (MF−LF, HF−LF, HF−MF) in PLC and OT separately and next conducted spatial correlation analyses. Under both treatments, higher levels of fear engaged widespread activation of brain regions involved in fear processing, with fear engaging distributed systems in the subcortical, limbic, prefrontal and default mode regions. Visual inspection indicated that under OT, less prefrontal regions were engaged, in particular in the high fear contexts (see HF−LF, HF−MF contrasts, **Fig. 2A**).

**Fig. 2.**
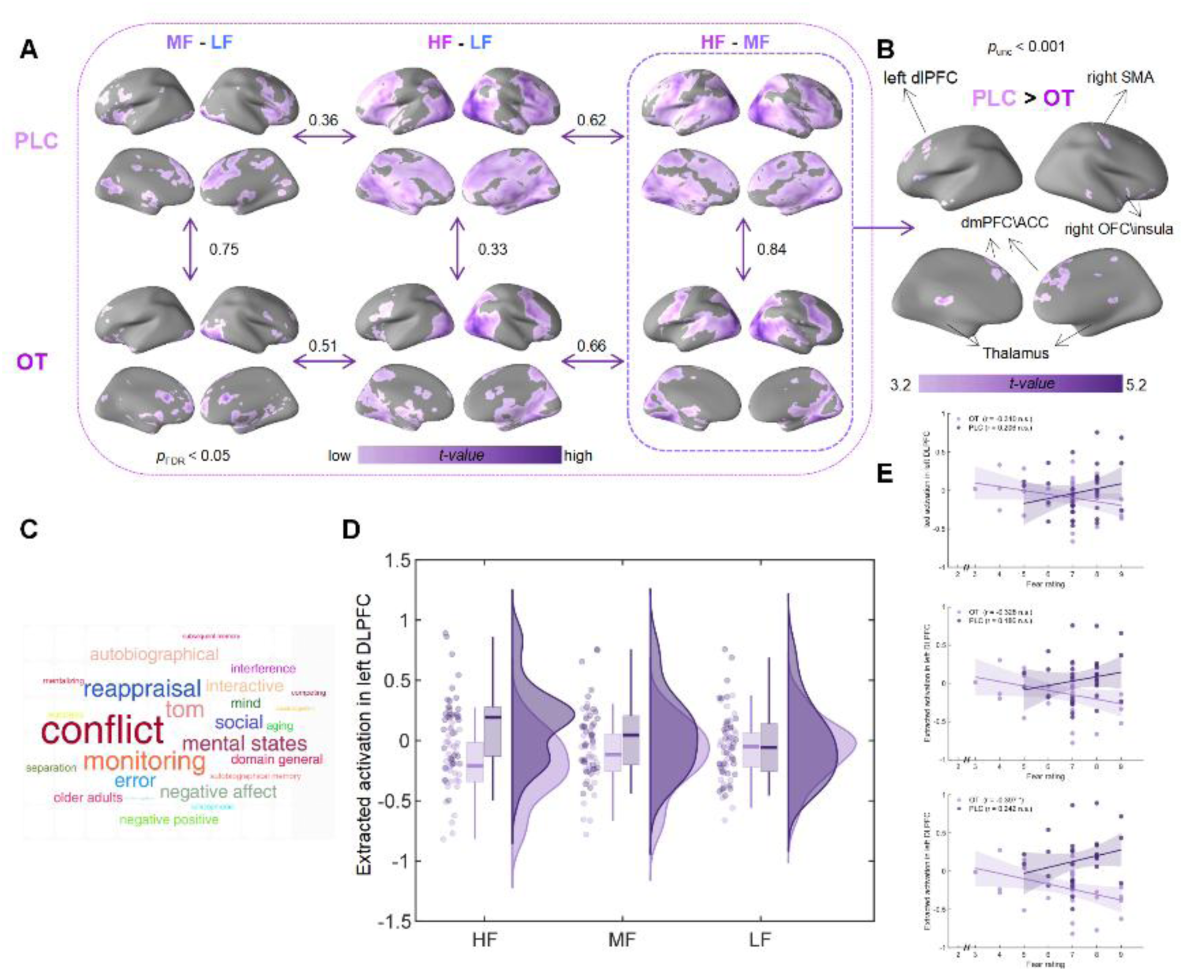
The effects of OT on dPFC activity and its association with subjective fear experience. **A** Neural representations under three intensities of fear in PLC and OT, and spatial correlations across conditions and group. **B** OT but not PLC decreased the activation in left dlPFC. **C** Fuctional decoding using NeuroSynth. **D** Parameter estimates extracted from the significant left dlPFC cluster are shown for descriptive visualization only. **E** Exploratory associations between dlPFC parameter estimates and post-exposure subjective fear. Abbreviations: OT oxytocin, PLC placebo, LF low fear, MF medium fear, HF high fear. FDR false discovery rate, dlPFC dorsolateral prefrontal cortex, dmPFC dorsomedial prefrontal cortex, ACC anterior cingulate cortex, OFC orbitofrontal cortex, SMA sensorimotor area. * *p*<0.05, *** *p*<0.001

Directly comparing the two treatment groups using two sample t-tests based on first-level HF−MF contrast suggested treatment effects on the left dorsolateral prefrontal cortex, right SMA and OFC\insula, bilateral dmPFC and ACC, and bilateral thalamus (initial uncorrected statistical results were reported in **Table S1**), with effects in the left dlPFC surviving correction for multiple comparison (left dlPFC, peak MNI, x, y, z=-33, 14, 44, *t*_(1,65)_=5.19, k=318, *p*_FWE-peak_=0.028, **Fig. 2B**). Parameter estimates extracted from this cluster across LF, MF, and HF were used only for descriptive visualization of the condition-specific pattern underlying the corrected whole-brain effect and were not subjected to additional inferential tests (**Fig. 2D**).

Exploratory brain-behavior analyses were conducted to characterize the association between dlPFC parameter estimates and post-exposure subjective fear ratings. These analyses were treated as exploratory and were not used as independent evidence for the treatment effect. The OT group showed a negative association between dlPFC activity during HF and post-exposure subjective fear (*r*=-0.397, *p*=0.02, **Fig. 2F**), whereas the corresponding association in the PLC group was not significant (*r*=0.242, *p*=0.168, **Fig. 2F**). Fisher r-z transformations indicated that these correlation coefficients were significantly different between the OT and PLC groups (Z=-2.61, two-tailed *p*=0.0091).

### OT enhanced dlPFC-amygdala functional connectivity in high fear contexts

Examining treatment-induced functional connectivity changes in the dlPFC revealed that OT enhanced connectivity between this region with the basolateral amygdala under the HF− MF contrast (peak MNI, x, y, z = 30, -1, -31, *t*_(1,65)_ = 4.11, k = 33, *p*_FWE-peak(svc)_ = 0.047, **Fig. 3A**). Connectivity estimates across LF, MF, and HF were extracted only for descriptive visualization of the condition-specific pattern and were not subjected to additional inferential testing on the same effect (**Fig. 3C** and **Fig. 3D**).

**Fig. 3.**
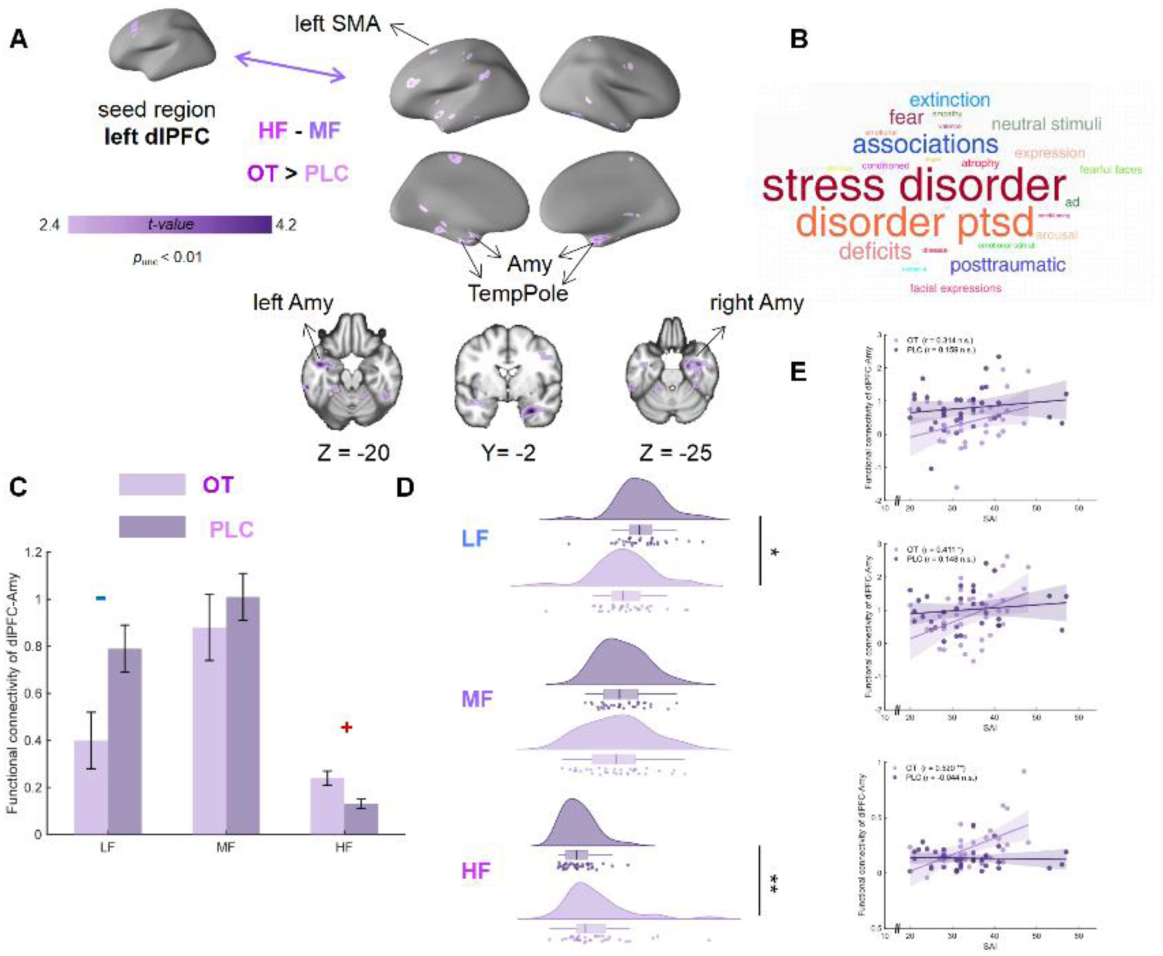
The effects of OT on left dPFC-Amygdala coupling and its association with subjective state anxiety. **A** OT enhanced functional connectivity between the left dlPFC and bilateral Amygdala (for display purpose, the threshold was set to *p*_unc_<0.01). **B** Fuctional decoding using NeuroSynth. **C** Trends of functional connectivity between left dlPFC and bilateral Amygdala in the two groups in response to changes of fear intensity. **D** Between-group comparisons of the functional connectivity between left dlPFC and bilateral Amygdala under different intensities of fear (for display purpose). **E** Exploratory associations between left dlPFC-Amygdala coupling and state anxiety. Abbreviations: OT oxytocin, PLC placebo, LF low fear, MF medium fear, HF high fear, dlPFC dorsolateral prefrontal cortex, Amy amygdala, TempPole temporal pole, SMA sensorimotor area. * *p*<0.05, ** *p*<0.01

Exploratory associations between dlPFC-amygdala coupling and state anxiety scores were examined to characterize potential behavioral relevance of the connectivity pattern. State anxiety was associated with dlPFC-amygdala connectivity in OT group (*r_MF_* = 0.411, *p_MF_* = 0.0175, *r_HF_*=0.520, *p_HF_*=0.0019, **Fig. 3E**) but not in PLC group (*r_MF_*=0.148, *p_MF_*=0.4045, *r_HF_*=-0.044, *p_HF_*=0.8049, **Fig. 3E**). Fisher r-z transformations indicated that correlation coefficients under HF were significantly different between the OT and PLC groups (Z=2.43, two-tailed *p*=0.015).

### OT strengthened FPN and DMN control over SCT and LN in high fear contexts

Examining the effects of OT on the large-scale network level using separate two-sample t-tests on the matrices of z-values for the three main contrasts of interest (MF−LF, HF−LF and HF−MF) demonstrated that compared to LF, OT significantly enhanced fuctional connectivity between FPN and SCT (*t*_(1,65)_ = 3.45, *p_FDR_* = 0.035), DMN and SCT (*t*_(1,65)_ = 2.98, *p* = 0.004), FPN and LN (*t*_(1,65)_=2.37, *p*=0.021), DMN and LN (*t*_(1,65)_=2.39, *p*=0.02) in HF **(Fig. 4B**). Similar but weaker functional connectivity patterns were observed under MF−LF contrast, DMN and LN (*t*_(1,65)_=2.12, *p*=0.038) as well as FPN and SCT (*t*_(1,65)_=2.06, *p*=0.043) showed OT effects **(Fig. 4B**). In contrast, the HF-MF contrast did not show this OT-related connectivity enhancement pattern **(Fig. 4B**). Consistent with these findings, Mantel tests comparing the OT-effect patterns across contrasts indicated that the network-level OT effects were highly similar between HF-LF and MF-LF (rho_HF-LF_MF-LF_=0.85, *p*<0.001), but not between HF-LF and HF-MF (rho_HF-LF_HF-MF_=0.03, *p*=0.459) or between MF-LF and HF-MF (rho_MF-LF_HF-MF_=-0.43, *p*=0.971) (**Fig. 4B**). Collectively, these results suggest that OT modulates large-scale functional connectivity in a consistent manner when comparing elevated fear (MF or HF) against LF (with stronger effects under HF-LF), whereas this network-level OT effect pattern is absent or qualitatively different when directly contrasting HF with MF (HF-MF).

**Fig. 4.**
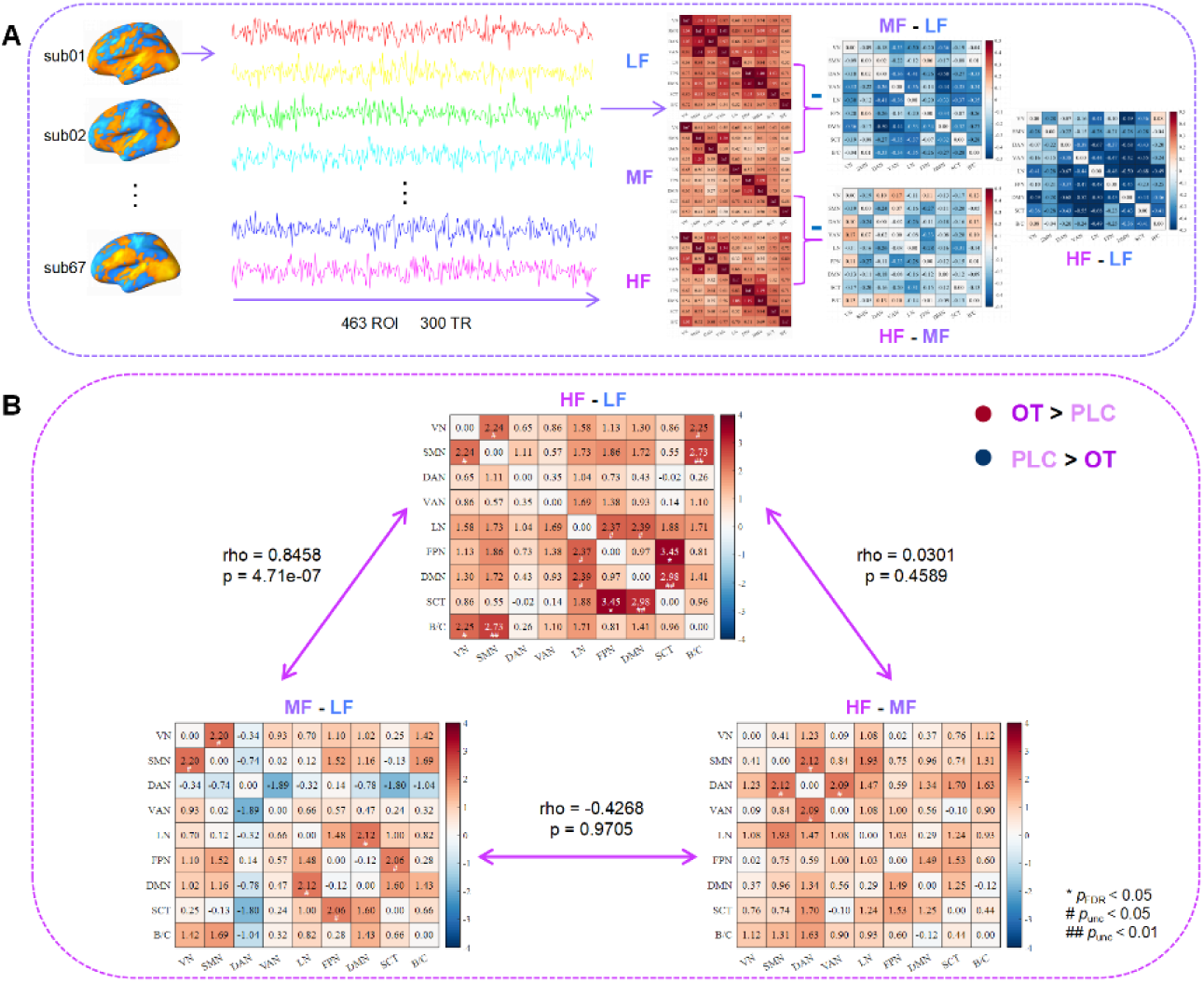
OT enhanced functional connectivity on the level of large-scale networks. **A** Flow of z-value matrix calculation for the degree of response functional connectivity under specific conditions and contrasts. **B** OT significantly strengthened the functional connectivity between FPN and SCT as well as LN, DMN and SCT as well as LN in the medium to high intensity of fear contexts compared to low fear condition. Abbreviations:OT oxytocin, PLC placebo, LF low fear, MF medium fear, HF high fear, VN visual network, SMN somatomotor network, DAN dorsal attention network, VAN ventral attention network, LN limbic network, FPN frontoparietal network, DMN default mode network, SCT subcortical network, B/C brainstem and cerebellum. * *p*_FDR_<0.05, ##*p*_un-corrected_<0.01, # *p*_un-corrected_<0.05.

### OT strengthened whole-brain distributed functional connectivity especially in the FPN and DMN or SCT and LN systems under high fear

Whole-brain functional connectivity analyses revealed a widely distributed OT-induced enhancement of connectivity linking the FPN and DMN with the SCT and LN, suggesting that OT strengthens communication between control-related networks (FPN and DMN) and fear expression–related networks (SCT and LN) when fear is elevated (MF and HF relative to LF). In contrast, this network-level OT effect pattern was not evident for the HF-MF contrast, indicating that OT does not similarly modulate connectivity for incremental differences between high and medium fear. Moreover, similarity analyses of the whole-brain OT-effect patterns showed that the HF-LF and MF-LF contrasts were more closely aligned than either was with the HF-MF contrast (rho_HF-LF_MF-LF_=0.11, *p*<0.001; rho_HF-LF_HF-MF_=0.02, p=0.009; rho_MF-LF_HF-MF_=-0.02, *p*=0.978, **Fig. 5**).

**Fig. 5.**
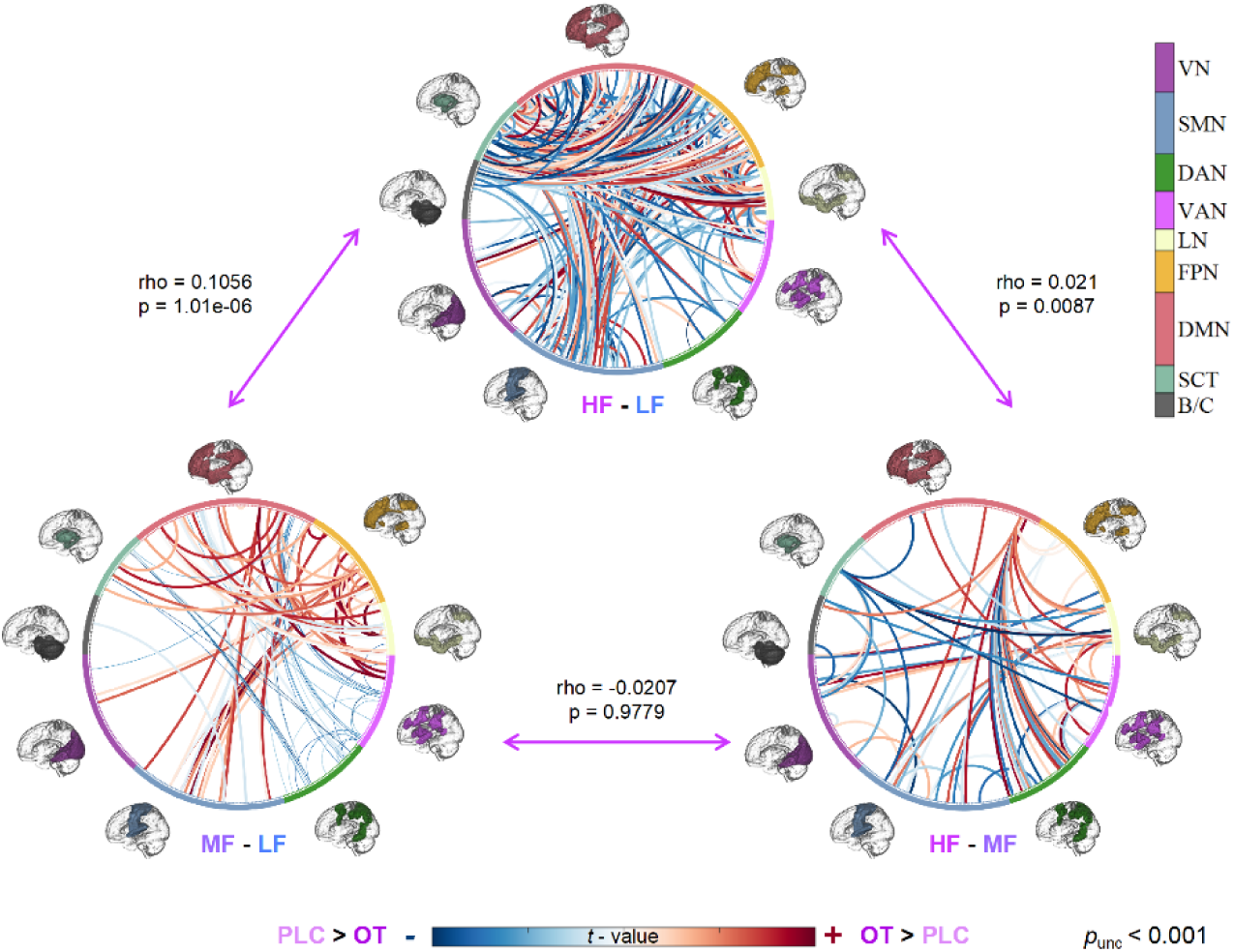
Whole-brain functional connectivity results. OT strengthened whole-brain distributed functional connectivity especially on FPN and DMN control on SCT and LN under medium to high fear contexts. Spatial correlation analyses demostrated that the whole-brain functional connectivity patterns under HF-LF and MF-LF contrasts are more similar than HF-MF contrast compared with the other two contrasts. Abbreviations: OT oxytocin, PLC placebo, LF low fear, MF medium fear, HF high fear, VN visual network, SMN somatomotor network, DAN dorsal attention network, VAN ventral attention network, LN limbic network, FPN frontoparietal network, DMN default mode network, SCT subcortical network, B/C brainstem and cerebellum.

### Treatment classification based on the response of a synergistic brain connectivity- and activity-based signature for subjective fear (CAFE)

To comprehensively determine and independently validate the fear-decreasing effects of OT, we utilized our recently developed neuromarker that accurately tracks subjective fear in naturalistic contexts by synergistically capitalizing on activity and connectivity^39^. In line with the univariate activation analysis and functional connectivity analysis results indicate that OT affects the neurofunctional signature of fear depending on the experienced fear intensity. The whole-brain MVPA results revealed that the fear signature could significant discriminate PLC and OT in the HF - MF contrast (mean ± SD of accuracy = 72 ± 5.5%, *p* = 0.0008, area under the curve (AUC) = 0.69, **Fig. 6 light purple pannel**), but not in the HF - LF contrast (mean ± SD of accuracy = 58 ± 6.0%, *p* = 1, AUC = 0.57, **Fig. 6 dark purple pannel**) and MF - LF contrast(mean ± SD of accuracy = 51 ± 6.1%, *p* = 0.2713, AUC = 0.43, **Fig. 6 pink pannel**).

**Fig. 6.**
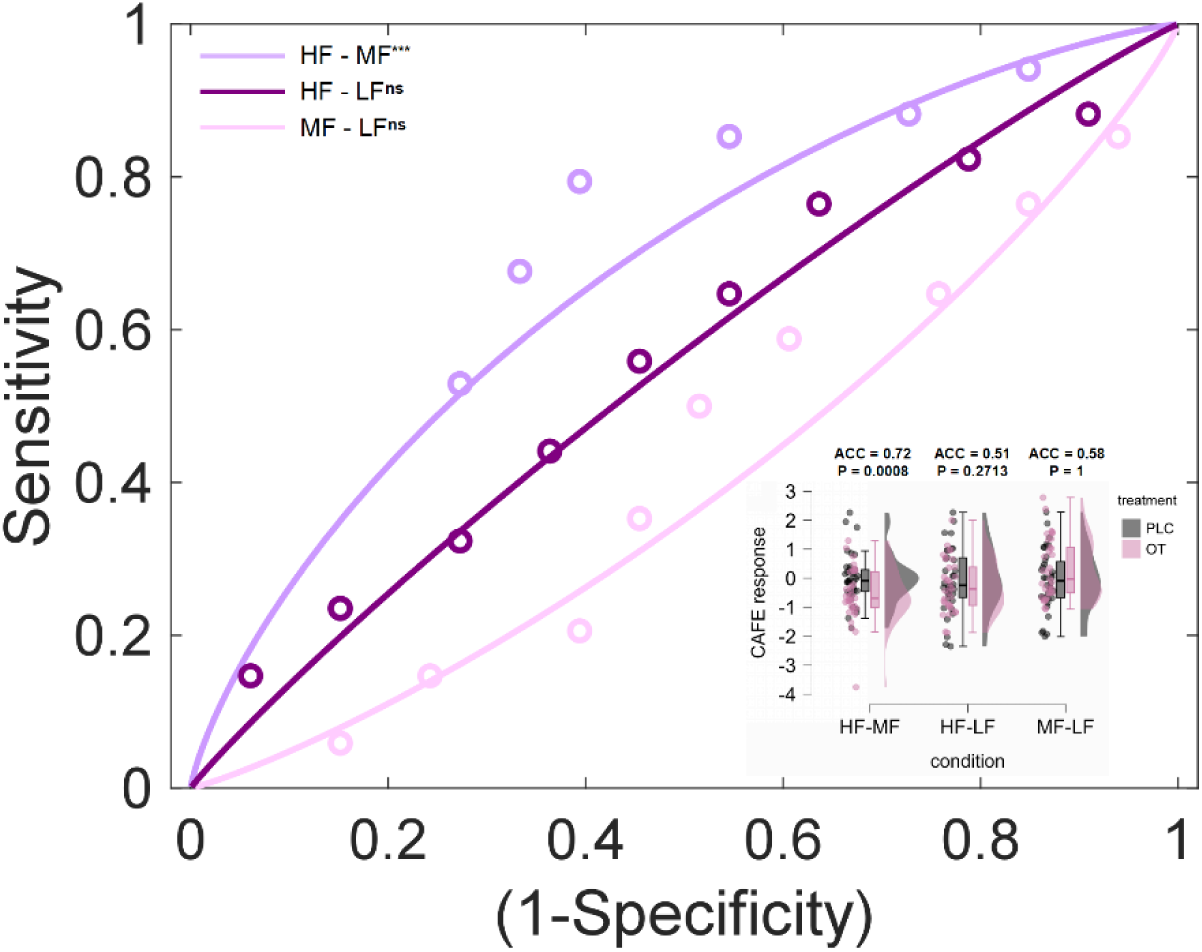
Treatment classification results based on CAFE response. CAFE successfully classify OT and PLC in high fear context, but not in extreme high fear and medium fear conditions. Abbreviations: OT oxytocin, PLC placebo, LF low fear, MF medium fear, HF high fear, ACC accuracy. ***<0.001, ns not significant

## Discussion

The present study combined the randomized placebo-controlled intranasal administration of OT with fcuntional neuroimaging in a naturalistic context that mimics the dynamic processing of fear in real-life situations with varying levels of fear to determine the acute behavioral and neural effects OT on the subjective experience of fear. Importantly, this work complements our previous study in which OT selectively reduced fear in social naturalistic contexts and modulated an lMCC–amygdala pathway alongside DAN-centered network communication^40^. Here, we extend that framework to an explicitly threatening dynamic setting and show that OT’s effects are most pronounced when fear is high, implicating a partly distinct control architecture.

At the behavioral level, OT specifically attenuates subjective fear following the threatening movie while the treatment groups did not show differences before the threat exposure, indicating that OT exerts its anxiolytic effects in a contexts-dependent manner. Further exploring the neural signatures of different levels of fear under naturalistic conditions revealed that OT exerted its neural effects particularly at higher levels of fear. OT significantly reduced engagement of a set of brain regions related to emotional experience and regulation, including the left dorsolateral prefrontal (dlPFC), bilateral dorsomedial prefrontal (dmPFC), and right insula, while it enhanced functional communication between left dlPFC and bilateral amygdala during periods of high subjective fear. Network analyses further demonstrated that – particulaley in high fear contexts - OT substantially strengthened functional connectivity between DMN and LN, FPN and SCT, and brain-wide coupling, potentially resulting in more accurate perception and control of excessive fear. Taken together, the results suggest that a single dose of intranasal OT has the potential to context-dependently reduce high fear in real-life situations by decreasing left dlPFC activation and enhancing its communication with the amygdala, while simultaneously affecting network-level interaction between regulatory (DMN and FPN) and subcortical (SCT and LN) systesms, as well as brain-wide functional communication, contributing to better regulatory control in highly fear-inducing situations.

Previous studies have suggested OT has a fear- and anxiety-reducing potential, yet results have been inconsistent and difficult to translate to complex fear situations that resemble fear in real-life contexts. Here, we combined naturalistic fMRI using a fear-inducing full-length film with a pre-registered, randomized, double-blind, placebo-controlled OT administration trial, reflecting increasing efforts to advance treatment evaluation via precision pharmco-neuroimaging under ecological valid conditions ^37,39,40,46,56^.

To enhance the ecological validity of treatment evaluation we utilized dynamic movie stimuli^37–39^ in combination with naturalistic fMRI with a fear-inducing whole-length movie and a pre-registered randomized double-blind placebo-controlled OT-administration trial. The procedure induced a highly immersive and comparably strong targeted emotional experience in pilot experiments and treatment studies^37–40^, while accounting for the dynamic nature of real-life emnotional experiences. OT – relative to PLC-effectively decreased subjective fear experiences following exposure to dynamic fear-inducing context. Importantly, the treatment groups did not differ before the threat exposure, suggesting that the anxiolytic effects of OT evolve in interaction with the context.

Although the naturalistic horror paradigm elicited a mixture of negative affective responses,^37,38^ the converging neural and behavioral findings suggest a relatively pronounced effect on fear. At the neural level, the HF–MF response showed stronger expression of the fear signature than signatures of anxiety and general negative affect, indicating that the observed neural pattern was more closely aligned with fear-related processing. Consistently, although the movie increased both arousal and escape motivation, OT did not significantly modulate these secondary behavioral measures, whereas its baseline-adjusted effect was evident for subjective fear. Together, these findings suggest that the observed OT effects were more closely associated with fear processing than with a generalized attenuation of negative affect, arousal, or avoidance motivation.

To further explore OT’s specific role in the dynamically changing fear contexts, we explored the effects of OT on neural activity at different fear levels. In terms of regional activation, OT was found to reduce the activation in the left dlPFC in high fear contexts. The dlPFC is a region with a well-established role in cognitive control and regulation^5,57–60^ and regulation of conditioned fear modulation^42,57,58^ while more recent accounts additionaly suggest a role in conscious emotional experiences, including the subjective experience of fear^2,4,61–63^. Exploratory Neurosynth decoding, now used only for functional interpretation, linked the identified region to terms related to conflict control, reappraisal, and negative affect. Furthermore, OT enhanced the functional connectivity between the left dlPFC and the bilateral amygdala. This pathway has been previously implicated in the top-down regulation of excessive fear-related amygdala engagement in fear-related disorders^64–66^. A further investigation of the brain-behaviour associations revealed that OT established a negative correlation between the activity of the left dlPFC and the level of subjective fear experiences, and a positive correlation between the functional coupling between the left dlPFC and the bilateral amygdala and the level of state anxiety (SAI). This evidence provides support for the behavioural relevance with the OT-induced neurofunctional changes. Our findings collectively indicate that OT exerts its regulatory effects on fear responses in highly naturalistic contexts by reducing the engagement of the dlPFC as regulatory and emotion representing regions while enhancing its control over the amygdala.

At the large-scale network level, OT markedly strengthened the control of cortical large-scale networks involved in emotion regulation (FPN and DMN) and subcortical systems (SCT, LN) in the high fear contexts. The FPN supports goal-directed action and cognitive control^67–69^, and the DMN promotes self-referential^70,71^ and social processing^72,73^, with recent evidence suggesting an additional role in emotional experience in dynamic naturalistic contexts^74–77^. As the sensory account of threat processing indicated^78^, threat-encoded sensory representations are relayed as threat-imbued sensory projections that terminate in major limbic areas (the amygdala, hippocampus, and insula) and (directly and indirectly) interact with various other cerebral and subcortical regions. Whereas previous research has demonstrated the effects of OT on the interplay between these large-scale networks during task-free conditions, these results highlight the behavioral relevance of these effects of OT in terms of the regulation of excessive subjective fear in high fear situations. Together with previous studies linking disruptions in these large-scale networks to emotional dysregulation in anxious and fearful disorders^42,70^, these findings suggest that OT may have therapeutic potential for related disorders^79,80^.

Network-level analyses, which examined the effects of OT at the fine-grained level of brain-wide connectivity patterns and employed a recently developed decoder that precisely tracks subjective fear under dynamic naturalistic conditions^39^, revealed that OT may broadly facilitate brain-wide information exchange within and across networks, particularly between higher-order FPN and DMN and lower-order SCT and LN in extreme fear contexts. The findings are consistent with the properties and effects of the distributed network-level OT signaling system^49,81,82^, as well as with recent findings suggesting that emotional processes such as fear and anxiety are processed in distributed networks and their interactions^4,39,45^. These findings may further underscore the therapeutic potential of the OT signaling system, along with the growing reconceptualization and increasing evidence that mental disorders are not related to dysfunction of a single brain region or pathway, but rather to dysregulated brain-wide communication^42,70,83^.

The findings need to be considered in the context of the limitations of the present design, including (1) to avoid sex and estradiol differences in the effects of OT^84–87^ on this study, we focused only on male participants, and future studies need to test generalization to females, (2) the study used a proof-of-concept design in healthy subjects, and future studies need to consolidate the effects of OT in clinical anxiety or PTSD populations, (3) although the naturalistic paradigm improves ecological validity, LF segments contained fewer TRs than MF and HF segments, which may reduce the stability of LF-related estimates. Moreover, fear intensity in naturalistic movies may covary with sensory, attentional, and narrative features. Future studies should model low-level audiovisual and narrative features explicitly to further disentangle fear intensity from other dynamic stimulus properties, (4) future studies are required to test the impact of repeated dosage of OT in naturalistic contexts to further validate its effects on fear reduction.^31,88^

In conclusion, the present study suggests that OT reduces post-exposure subjective fear in dynamic naturalistic contexts after accounting for baseline ratings by decreasing the involvement of the left dlPFC and strengthening its regulatory control over the amygdala, as well as improving functional communication between emotion-control-related large-scale networks and brain-wide communication. These findings provide preliminary evidence that OT can influence fear-related neural dynamics under ecologically valid conditions. Future studies in mixed-sex and clinical samples are required before drawing strong therapeutic conclusions for anxiety disorders, PTSD, or other fear-related conditions^11,32,34^, or may help to prevent the development of these disorders during early stages or promote long-term recovery^89^ via improving the quality of life, and reduce the social and economic burden related to fear-related disorders.

## Methods

### Participants

Sixty-nine right-handed male participants with normal or corrected-normal vision were recruited from the University of Electronic Science and Technology of China. The sample size was originally determined based on the broader preregistered experimental protocol. To specifically evaluate the statistical power of the design examined in the present study, we additionally performed a G*Power analysis (v3.1) based on a 2 × 2 mixed design, with treatment (OT vs. PLC) as the between-subject factor and session (baseline vs. Long Fear) as the within-subject factor. Assuming a medium effect size (*f* = 0.25), *α* = 0.05, and 95% power, the analysis indicated a required total sample size of 54 participants. To reduce variance related to sex differences in the effects of OT^85,90^, the present study focused only on male subjects and adhered to validated exclusion criteria (details see supplementary methods). The final sample size was n = 67 (mean ± SD, age = 21.03 ± 1.95 years), data from two participants were excluded (withdraw due to personal reasons, technical MRI issues). All participants provided written informed consent. The experimental protocol was prospectively registered on Clinical Trials.gov (https://clinicaltrials.gov/NCT05896553), approved by the ethics committee at the University of Electronic Science and Technology of China (UESTC-1061423041725893) and in line with the latest Declaration of Helsinki.

Using a randomized, double-blind, placebo-controlled, parallel-group pharmacological fMRI design, participants were randomly assigned to receive either a single intranasal dose of oxytocin (OT, 24IU-Sichuan Defeng Pharmaceutical Co. Ltd, China) or placebo (PLC, same ingredients i.e. sodium chloride and glycerin but without the peptide). The spray bottles were identical and dispensed by an independent researcher based on a computer-generated randomization sequence to ensure double-blinding. The investigators involved in data acquisition and analyses were blinded for group allocation. In line with recent recommendations, the fMRI assessment began 45 minutes after treatment administration^91,92^. The Long Fear task analyzed in the present study was the second task in the scanning session and began approximately 80 min after treatment administration. It was preceded by a shorter naturalistic fear task examining oxytocin effects on social versus non-social fear processing, which was independently preregistered and has been reported previously.^40^ Accumulating pharmacokinetic and neuroimaging evidence also indicates that our sampling interval of 70-90 min falls within the period of OT’s central effects.^93–95^

To control for pre-treatment between-group differences, participants completed a series of mood and mental health questionnaires (as detailed in **Table 1**). Participants were asked to guess which treatment they had received after the experiment (with a non-significant treatment guess *χ*^2^=0.39, *p*=0.53, confirming successful double-blinding). Additionally, participants were required to complete a surprise recognition task after the experiment that involved presenting stills of the horror movie shown during fMRI and new scenes to test the attentive processing of the stimuli. All participants achieved a re-recognition accuracy of over 80% and no significant difference in performance was observed between the two treatment groups (*t*_(1,65)_=-0.244, *p*= 0.808).

### Naturalistic dynamic fear paradigm using long movies

The task began 80 minutes after the treatment administration (**Fig. 1A & Fig. 1B**). During the naturalistic fear movie paradigm participants were instructed to watch a 10-min-long horror movie with soundtrack attentively. To quantify the influence of long horror naturalistic contexts and OT’s treatment effects on subjective fear, the participants were asked to rate their level of fear experience using a 9-point Likert scale, with 1 indicating no fear and 9 indicating very strong fear before the first rest-fMRI scanning (as baseline) and after the task (**Fig. 1D**).

Fear intensity during the movie was determined using continuous ratings obtained from an independent sample (N=20, 10 males, age=24.2±2.1). Rather than providing discrete ratings for each fMRI volume, participants continuously tracked their moment-to-moment fear experience using a mouse-position rating procedure while watching the 10-min horror movie. This procedure yielded 1187 time-stamped rating samples across the movie. To match the fMRI temporal resolution, the continuous rating time series was downsampled to one value every 2 s, corresponding to the TR of the fMRI acquisition. Each TR was then classified into low-, medium-, or high-fear intensity according to the downsampled rating value. Based on these ratings the entire horror movie was split into segments corresponding to low (rating 0-30, 44 TR), medium (rating 31-60, 132 TR) and high (ratings 61-90, 124 TR) levels of fear (LF\MF\HF) (**Fig. 1C**). The tasks were programmed in Python 3.7 using PsychoPy (version 2022.2.4).

### MRI data acquisition and preprocessing

MRI data were collected on a 3.0-T GE Discovery MR750 system (General Electric Medical System, Milwaukee, WI, USA) and preprocessed using FMRIPREP 21.0.0, a Nipype 1.6.1 based tool that integrates preprocessing routines from different software packages and SPM 12 (Statistical Parametric Mapping; http://www.fil.ion.ucl.ac.uk/spm/; Wellcome Trust Centre for Neuroimaging, detailed in supplementary methods).

Next, a voxel-wise general linear model (GLM) was conducted for each participant. Specifically, a first-level model was designed that included separate regressors for the three levels of fear intensity (LF\MF\HF). In line with previous studies, we removed variance associated with the mean, linear and quadratic trends, the average signal within anatomically-derived CSF mask, the effects of motion estimated during the head-motion correction using an expanded set of 24 motion parameters (six realignment parameters, their squares, their derivatives, and their squared derivatives) and motion spikes (FMRIPREP default: FD>0.5mm or standardized DVARS>1.5).

### Behavioral analysis of subjective fear ratings

Subjective fear ratings were analyzed to examine OT effects on post-exposure fear following the Long Fear movie. First, a mixed repeated-measures ANOVA was conducted with timepoint (pre-exposure baseline vs. post-exposure Long Fear) as a within-subject factor and treatment group (oxytocin vs. placebo) as a between-subject factor. This analysis tested the main effects of timepoint and treatment as well as their interaction. Partial eta-squared (ηp²) was reported as the effect size.

Because the primary behavioral question concerned treatment effects on fear following threat exposure while accounting for pre-exposure ratings, we further conducted an ANCOVA with post-exposure Long Fear rating as the dependent variable, treatment group as the between-subject factor, and pre-exposure baseline rating as a covariate. The homogeneity of regression slopes assumption was evaluated by testing the baseline × treatment interaction before fitting the final ANCOVA model. Adjusted group means and 95% confidence intervals were estimated at the grand mean of baseline ratings.

To examine whether the behavioral effect of OT reflected subjective fear rather than a broader change in general emotional arousal or avoidance-related motivation, the same mixed ANOVA and baseline-adjusted ANCOVA framework was additionally applied to arousal and escape motivation ratings. These analyses are reported in the Supplementary Materials.

Exploratory follow-up comparisons were conducted to characterize the behavioral pattern. Independent-samples t-tests were used to compare treatment groups at each timepoint, and paired-samples t-tests were used to assess within-group changes from baseline to the Long Fear phase. Change scores, defined as Long Fear minus baseline ratings, were additionally compared between groups. All tests were two-tailed, and significance was set at p < 0.05.

### Effects of OT on fear and emotion regulation related brain activity

To examine the effects of OT and its interactions with fear intensity a GLM-based model was established in SPM 12. The model incorporated three regressors for the fear levels: LF, MF, HF. The 24 head motion parameters were included as covariates of no interest. Key first-level contrasts of interest were contrasts that modeled fear intensity related differences in neural activity (HF−LF, MF−LF and HF−MF). Among these contrasts, HF > MF was defined a priori as the primary planned contrast of interest because it directly tested whether OT modulated neural responses to high fear relative to a lower but still fear-relevant intensity condition. Importantly, MF and HF segments were expected to be more closely matched in general perceptual input, salience, attentional engagement, and narrative involvement, thereby providing a more targeted comparison of fear intensity within fear-eliciting contexts. In contrast, LF segments may have included more heterogeneous low-threat or transitional scenes and also contained substantially fewer time points than MF and HF. Therefore, HF > MF was considered the most appropriate primary contrast for assessing intensity-dependent fear modulation by OT. Specifically, We first computed the spatial similarity between the two treatment groups (OT and PLC) and the three fear levels (MF−LF, HF−LF and HF−MF) using pattern similarity module in CanlabCore (https://github.com/canlab/CanlabCore/blob/master/CanlabCore/Statistics_tools/canlab_pattern_similarity.m). At the second level, voxel-wise independent-samples t-tests were performed to directly compare the OT and PLC groups using the first-level contrast images representing the HF–MF effect as input. All resulting maps from the whole brain analyses were thresholded at the peak level of family-wise error (FWE) correction at *p* < 0.05^96^.

To visualize the condition-specific pattern underlying significant whole-brain effects, parameter estimates were extracted from significant cluster (left dlPFC, MNI = -30, 17, 40) for the LF, MF, and HF conditions. These extracted values were used only for descriptive visualization of the direction and pattern of the effect across fear intensity levels. No additional inferential statistics were conducted on these extracted values for the same effect, thereby avoiding circular inference. Correlation analysis was conducted to determine associations between the behavioral (subjective fear underwent horror movie) and neural effects of OT (both activation and functional connectivity between specific brain regions) using Graghpad Prism.

Independent meta-analytic functional decoding of significant whole-brain clusters was conducted using Neurosynth to provide an exploratory functional characterization of the identified regions. This decoding analysis was used for interpretation only and was not treated as confirmatory evidence for the treatment effect. Specifically, after thresholding the activation t-value map in response to OT (*p_peak-level_*<0.001, k>20, **Fig. 2B**), we used Neurosynth to decode the functions after saving it as separate nii file. Word clouds were plotted based on the decoded functions in the top 25 positions (excluded structural words).

### Effects of OT on functional connectivity of the regions identified

We further examined the regions that exhibited effects of OT on th level of neural activity changed their functional communication with other regions using a seed-to-whole-brain general Psychophysiological interaction (gPPI) analysis. gPPI examines functional connectivity between brain regions that is contingent on a psychological context. We used the Generalized PPI toolbox as implemented in SPM 12, to conduct gPPI. This method includes additional task regressors to reduce the likelihood that the functional connectivity estimates are driven simply by co-activation and has been widely used in similar studies. Based on our finding that OT decreased activation in left dlPFC under the contrast of HF−MF, we therefore computed the functional communication of this region (using a 6mm radius which centered in MNI = -30, 17, 40) with other voxels in the whole brain under all conditions and contrasts, and two-sample t-tests were conducted to confirm the treatment effect of OT. Our hypothesis focuses on the control of the superior dlPFC over the inferior evolutionarily conserved amygdala, separate atlas-derived bilateral masks for the amygdala(independent centromedial amygdala and basolateral amygdala) were used for SVC analyses with FWE correction applied at peak level.

Connectivity estimates from significant gPPI clusters were extracted only for descriptive visualization of the condition-specific pattern. No additional inferential statistics were conducted on these extracted values for the same effect, avoiding circular inference.

### Effects of OT on large-scale brain network functional connectivity

Given that several studies reported that fear in naturalistic contexts requires a distributed functional communication on the network level and that OT exerts network level effects^40,42,97,98^, we examined whether OT modulates large scale network dynamics and interactions. In line with our previous study^39,40^, time-series of 463 atlas based regions-of-interest (ROIs) representing the whole brain were extracted (see **supplementary methods**), resulting in 463 BOLD time series of the size of *r*×*t* (*r* was the number of ROIs and *t* was the BOLD time series length of the whole long horror movie run) for each subject. The independently determined three levels of fear (HF\MF\LF) were included with the onsets and duration shifted by 3 TRs to account for the hemodynamic lag. Based on the temporal information of the three fear levels, three separate *r*×*d* matrices (where *r* is the number of ROIs and *d* is the BOLD time series length of each condition) were constructed for HF, MF, LF.

A matrix of size *r*×*r* was obtained by calculating Pearson’s correlation between the average BOLD time series of each pair of ROIs for each condition. Additionally, Fisher’s z-transform was applied to improve the normality of the correlation coefficients. For the subsequent analyses the data was examined on the level of large scale brain networks (9 x 9, definition of network, please see **supplementary methods**) and individual brain regions (463×463) based on Z-value matrics for each fear level.

It is important to note that r values are not additive or subtractive, but Z scores are summative. Matrix of Z-values corresponding to particular contrasts (e.g. Z_HF-LF_) can be computed by subtract the matrices of Z-values corresponding to different conditions (e.g. Z_HF_-Z_LF_). Therefore, for each participant the subtracted Z-values matrix under specific contrasts (Z_HF-LF_\Z_MF-LF_\Z_HF-MF_) were computed. We then conducted two-sample t-tests on each pair of network-level functional connectivity to determine treatment effects. In addition, spatial similarity between neural matrices was assessed using a permutation-based matrix similarity analysis^99,100^. Specifically, the corresponding upper-triangular elements of two symmetric neural similarity matrices were correlated, yielding a similarity coefficient reported as rho, and statistical significance was assessed using permutation testing. This procedure follows the logic of Mantel-test-based matrix comparison^101^ and second-order representational similarity analysis, which has been used to quantify representational relationships or representational connectivity between neural patterns, brain regions, and experimental conditions.

### Effects of OT on whole-brain functional connectivity

To further investigate the impact of OT on a finer spatial scale, we analyzed whole-brain functional connectivity using two-sample t-tests on the Z-value matrices for the main contrasts of interest (HF-LF, MF-LF and HF-MF). Mantel tests were performed on the between-group whole-brain functional connectivity matrices between the three key contrasts to assess their similarity.

### Effects of OT on synergistic brain connectivity- and activity-based signature for subjective fear (CAFE)

We applied a recently developed whole-brain multivariate neuromarker for fear in naturalistic contexts which synergistically capitalizes on brain connectivity- and activity-based signature (CAFE^39^) to accurately track subjective fear in dynamic naturalistic contexts to independently validate effects of OT on the whole-brain expression of different intensities of fear. First, we calculated the functional connectivity matrix (463×463) for each subject under the three conditions (HF\MF\LF) and applied dot-product the integrated one-dimensional vector (n = 107253 features in total) with the CAFE weights to calculate the CAFE response for each subject under each of the three conditions. Second, in line with the other analyses we build the contrasts of interest (HF-LF, MF-LF and HF-MF) and calculated CAFE response of each contrast. To quantify the classification performance of the model (classification performance between PLC and OT under same contrast condition), Receiver Operating Characteristic (ROC) analysis was conducted.

## Supporting information

SI for OT fear study

## Data Availability

All data produced in the present study are available upon reasonable request to the authors.

## Acknowledgement

This work was supported by the China MOST2030 Brain Project (Grant No. 2022ZD0208500-DY), the UGC General Research Fund (Grant No. 17615525-BB), and a seed start-up grant from The University of Hong Kong, including a collaborative project seed grant (Grant No. 17615525-17615525).

## Author contributions

KF and BB1 designed the study. KF, ZZ and DL conducted the experiment and collected the data. KF, SX, WZ, DY performed the data analysis with the help of QL, JH, TX, CL, JW, YZ, XZ, CL, MH, ML, ZL and BB2. KF and BB1 wrote the manuscript. SX, FZ, KK, TX, YZ, DY, and WZ critically revised the manuscript draft.

Note: BB^1^, Benjamin Becker; BB^2^, Bharat Biswal.

