## Supplementary material for "Oxytocin reshapes fear control: intensity-dependent prefrontal dominance and whole-brain integration during naturalistic viewing": SI for OT fear study

#### Supplemental Methods

##### Participants

Exclusion criteria of participants comprised a history of head injury, color blindness, current or regular substance or medication use, current or history of medical or psychiatric disorders, and any contraindications for oxytocin or MRI.

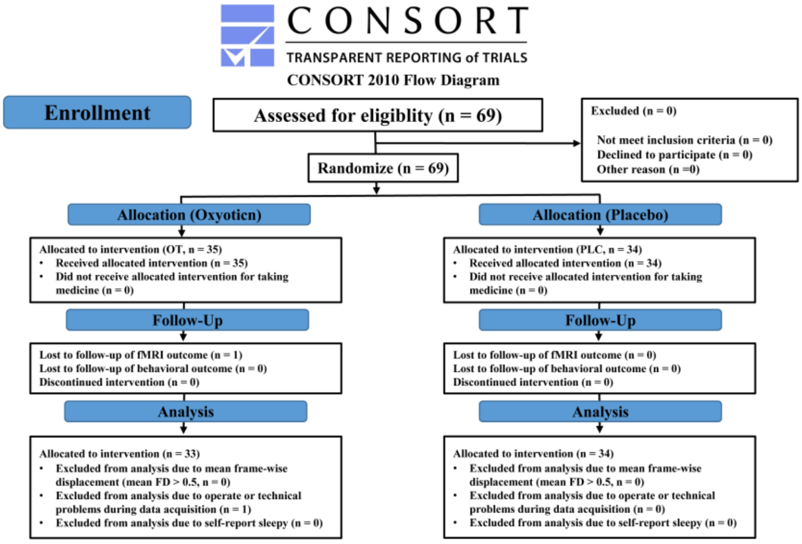


**Fig. S1 CONSORT Flow Diagram.**

##### Supplementary behavioral analyses of arousal and escape motivation

To assess whether the OT effect on subjective fear reflected a broader modulation of negative affective arousal or avoidance-related motivation, we conducted parallel supplementary analyses for arousal and escape motivation ratings. For each rating dimension, a mixed repeated-measures ANOVA was conducted with timepoint (pre-exposure baseline vs. post-exposure Long Fear) as a within-subject factor and treatment group (oxytocin vs. placebo) as a between-subject factor. In addition, baseline-adjusted ANCOVAs were conducted with post-exposure arousal or escape motivation rating as the dependent variable, treatment group as the between-subject factor, and the corresponding pre-exposure baseline rating as a covariate. The homogeneity of regression slopes assumption was evaluated before fitting each final ANCOVA model. These analyses were used to determine whether OT effects generalized to broader affective arousal or escape motivation, or were more selectively observed for subjective fear.

#### MRI acquisition and preprocessing

Functional MRI data was acquired using a T2*-weighted echo-planar imaging (EPI) pulse sequence (repetition time = 2000 ms, echo time = 30 ms, 36 slices, slice thickness = 3.8 mm, no gap, field of view = 200 × 200 mm, resolution = 64 × 64, flip angle = 90°, 3.125 × 3.125 × 3.8 mm voxels, axial scan plane). To improve spatial normalization and exclude participants with apparent brain pathologies a high-resolution T1-weighted image was acquired using a 3D spoiled gradient recalled (SPGR) sequence (176 slices, repetition time = 8.22 ms, echo time = 3.15 ms, field of view = 256 × 256 mm, resolution = 256 × 256, flip angle = 8°, 1 × 1 × 1 mm voxels).

The MRI images preprocessing included 1) Spatial normalization to the ICBM 152 Nonlinear Asymmetrical template version 2009c was performed through nonlinear registration with the antsRegistration tool of ANTs v2.3.3 [1], using brain-extracted versions of both T1w volume and template. 2) Brain tissue segmentation of cerebrospinal fluid (CSF), white matter (WM) and gray matter (GM) was performed on the brain-extracted T1w using FAST (FSL v6.0.5.1) [2]. 3) Before the automated preprocessing, 5 initial volumes of fMRI data were removed to allow for image intensity stabilization. 4) Next, functional data was slice time corrected using 3dTshift from AFNI [3] and motion corrected using mcflirt (FSL v6.0.5.1). This was followed by 5) co-registration to the corresponding T1w using boundary-based registration [4] with six degrees of freedom, using FLIRT (FSL). 6) Motion correcting transformations, 7) BOLD-to-T1w transformation and 8) T1w-to-template (MNI) warp (interpolated to 2 mm isotropic voxels) were concatenated and applied in a single step using ants ApplyTransforms (ANTs v2.3.3) using Lanczos interpolation.

Preprocessed images were spatially smoothed using an 8-mm full-width at half maximum (FWHM) Gaussian kernel in SPM 12.

#### Definiton of ROIs

The template utilized in this study comprises 400 cortical regions from the Schaefer atlas [5], 34 subcortical regions from the Melbourne subcortex atlas [6], and the reinforcement learning atlas (extended amygdala and hypothalamus) [7] and periaqueductal gray (PAG), brainstem, and cerebellar regions (n = 29) [8], resulting in a total of 463 regions. For large-scale network-level analyses, the 463 regions were assigned to nine whole-brain networks. The 400 cortical parcels from the Schaefer atlas were labeled according to the Yeo 7-network organization: Visual Network, Somatomotor Network, Dorsal Attention Network, Ventral Attention Network, Limbic Network, Frontoparietal Network, and Default Mode Network. The subcortical regions from the Melbourne subcortex atlas, together with the extended amygdala, hypothalamus, and periaqueductal gray regions, were grouped as a Subcortical Network. Brainstem and cerebellar regions were grouped as a Brainstem/Cerebellum network. This resulted in nine large-scale networks covering cortical, subcortical, brainstem, and cerebellar systems and insistent with previous work.[9, 10]

#### External Signature Validation

To validate the affective specificity of the HF-MF contrast, subject-level contrast maps were projected onto three independent external signatures: subjective fear (visual induced fear signature, VIFS)[11], anxiety (shock uncertainty-induced threat anticipation signature, SUITAS)[12], and general negative affect (picture induced negative emotion signature, PINES)[13]. For each participant, pattern expression was computed as a normalized dot product between the contrast map and each signature:

$$\text{expression} = \text{sum}(\text{beta} \times \text{weight}) / \text{sqrt}(\text{sum}(\text{weight}^2))$$

Group-level expression was tested against zero using one-sample t-tests and sign-flip permutation tests. Fear specificity was assessed by comparing fear expression with anxiety and PINES expression using paired one-tailed sign-flip permutation tests, followed by FDR correction. As supplementary classification analyses, each signature was used to discriminate HF-MF from the sign-reversed MF-HF contrast. AUC and zero-threshold accuracy were calculated for each signature. Pairwise permutation tests compared whether the fear signature showed higher AUC or accuracy than anxiety and PINES, with FDR correction across comparisons. At last, spatial similarity among external signatures was assessed using voxelwise Pearson correlation across gray-matter voxels. To further quantify overlap among the most strongly weighted voxels, each signature was binarized by retaining the top 5% absolute-weight voxels, and pairwise overlap was quantified using the Dice coefficient.

### Supplemental Results

##### Comparable arousal and escape motivation ratings between groups

To examine whether the behavioral effect of OT on subjective fear reflected a broader modulation of affective arousal or avoidance-related motivation, we applied the same analytical framework to arousal and escape motivation ratings.

For escape motivation, the mixed repeated-measures ANOVA revealed a significant main effect of timepoint ($F_{(1,65)} = 179.19$, $p < 0.0001$, $\eta_p^2 = 0.734$), indicating that the Long Fear movie robustly increased escape motivation across participants. However, neither the main effect of treatment ($F_{(1,65)} = 0.86$, $p = 0.358$, $\eta_p^2 = 0.013$), nor the treatment × timepoint interaction ($F(1,65) = 1.23$, $p = 0.272$, $\eta_p^2 = 0.019$), was significant. In the baseline-adjusted ANCOVA, the homogeneity of regression slopes assumption was satisfied, $p = 0.361$. Baseline escape motivation significantly predicted post-exposure escape motivation, $\beta = 0.276$, SE = 0.122, $t_{(64)} = 2.26$, $p = 0.027$, $\eta_p^2 = 0.074$. However, the treatment effect was not significant, $\beta = 0.130$, SE = 0.406, $t_{(64)} = 0.32$, $p = 0.749$, $\eta_p^2 = 0.002$. The adjusted post-exposure escape motivation means were 6.98 for the PLC group and 7.11 for the OT group.

For arousal ratings, the mixed repeated-measures ANOVA similarly revealed a significant main effect of timepoint, $F_{(1,65)} = 213.43$, $p < 0.0001$, $\eta_p^2 = 0.767$, indicating that the Long Fear movie strongly increased subjective arousal. The main effect of treatment did not reach significance, $F_{(1,65)} = 3.47$, $p = 0.067$, $\eta_p^2 = 0.051$, and the treatment × timepoint interaction was also not significant, $F_{(1,65)} = 0.72$, $p = 0.400$, $\eta_p^2 = 0.011$. The baseline × treatment interaction in the homogeneity of regression slopes test was significant, $p = 0.027$, indicating that the standard ANCOVA assumption was not fully met for arousal ratings. Therefore, the baseline-adjusted arousal ANCOVA should be interpreted cautiously. Nevertheless, the main-effects ANCOVA did not indicate a significant treatment effect on post-exposure arousal, $\beta = -0.169$, SE = 0.331, $t_{(64)} = -0.51$, $p = 0.611$, $\eta_p^2 = 0.004$.

Together, these results indicate that the Long Fear movie robustly increased both arousal and escape motivation, confirming the affective impact of the naturalistic threat exposure. However, OT did not significantly reduce post-exposure arousal or escape motivation. These findings suggest that the baseline-adjusted behavioral effect observed in the main analysis was more closely related to subjective fear than to a generalized reduction in affective arousal or avoidance-related motivation.

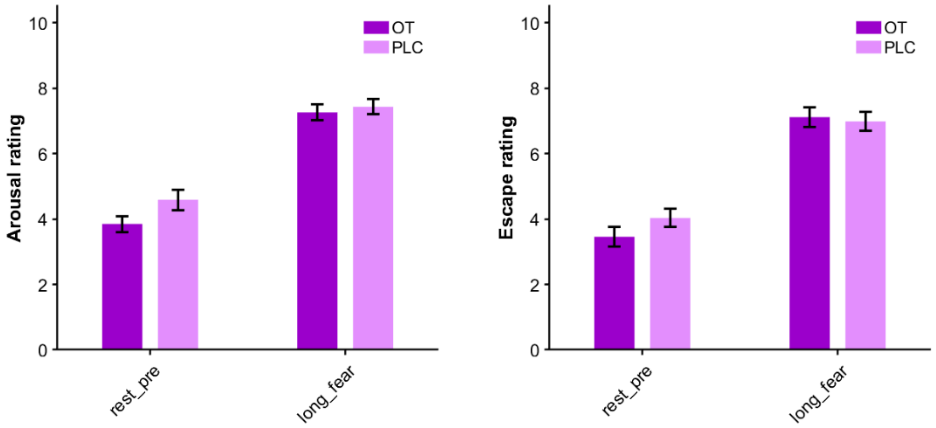


**Fig. S2 Comparable arousal and escape ratings across session**

**Fear-dominant external signature expression**

All three external signatures showed significant positive expression in the HF-MF contrast: fear (VIFS), $t_{(66)} = 7.82$, $p < .001$, q < .001, Cohen's $d = 0.96$; anxiety, $t_{(66)} = 6.30$, $p < .001$, q < .001, $d = 0.77$; and PINES, $t_{(66)} = 6.79$, $p < .001$, q < .001, $d = 0.83$. Importantly, fear-pattern expression was significantly stronger than both anxiety expression, Δ = 1.40, sign-flip p = .002, FDR-corrected q = .002, and PINES expression, Δ = 1.52, sign-flip p = .001, q = .002.

Supplementary classification analyses showed that all three signatures significantly discriminated HF-MF from the sign-reversed MF-HF contrast, all AUCs ≥ 0.86 and all qs < .001. However, pairwise permutation tests did not show significantly higher AUC for fear than anxiety or PINES after correction. For zero-threshold accuracy, fear showed a nominal advantage over anxiety, ΔACC = 0.134, p = .032, but this did not survive FDR correction, q = .065. Thus, fear specificity was mainly reflected in stronger pattern expression rather than superior classification performance.

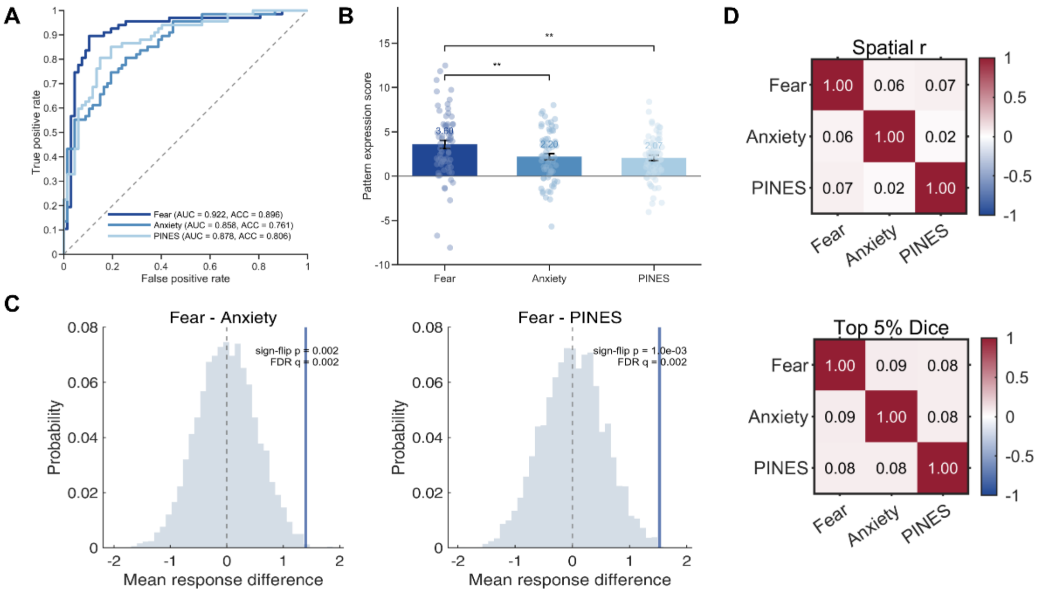


**Fig S3 Validation and specificity of fear-related neural pattern expression**

#### OT decreased widespread brain activation in high fear

**Table. S1 Initial OT-driven brain activation results in high fear**

| brain region | MNI(x,y,z) | cluster level | | | peak level | | | |
|---|---|---|---|---|---|---|---|---|
| | | $p_{FWE\text{-}corr}$ | $P_{uncorr}$ | k | $p_{FWE\text{-}corr}$ | $P_{uncorr}$ | $t$ | $Z$ |
| LH-dlPFC | -33, 14, 44 | 0.055 | 0.008 | 318 | 0.028 | 0.000 | 5.19 | 4.73 |
| RH-ACC | 14, 38, 30 | 0.000 | 0.000 | 1499 | 0.163 | 0.480 | 4.60 | 4.27 |
| extending to SFG | | | | | | | | |
| RH-OFC | 38, 28, -25 | 0.033 | 0.005 | 373 | 0.224 | 0.000 | 4.49 | 4.17 |
| LH-PCC | -5, -37, 8 | 0.339 | 0.061 | 142 | 0.353 | 0.000 | 4.30 | 4.02 |
| LH-SMA | -3, 28, 58 | 0.051 | 0.008 | 326 | 0.481 | 0.000 | 4.15 | 3.90 |
| extending to SFG | | | | | | | | |
| LH-cerebellum | -49, -59, -53 | 0.904 | 0.348 | 33 | 0.488 | 0.000 | 4.15 | 3.89 |
| RH-paracentral lobule | 14, -35, 58 | 0.858 | 0.290 | 42 | 0.545 | 0.000 | 4.09 | 3.84 |
| extending to precuneus | | | | | | | | |
| LH-IFG | -47, 28, 6 | 0.841 | 0.274 | 45 | 0.669 | 0.000 | 3.96 | 3.74 |
| RH-precentral | 34, -17, 44 | 0.332 | 0.060 | 144 | 0.732 | 0.000 | 3.90 | 3.68 |
| RH-insula | 40, 12, -5 | 0.931 | 0.398 | 27 | 0.874 | 0.000 | 3.72 | 3.53 |

$p_{FWE\text{-}corr}$ represented thresholds results $p < 0.05$(FWE Multiple Comparison Correction), and for display purpose only cluster size $k > 25$ was listed here. FWE, family wise error.

2. Zhang Y, Brady M, Smith S. Segmentation of brain MR images through a hidden Markov random

field model and the expectation-maximization algorithm. *IEEE Transactions on Medical Imaging* 2001; **20**(1)**:** 45-57.
